# Approaches used to enhance transition and retention for newly qualified nurses (NQNs): a rapid evidence assessment

**DOI:** 10.1101/2020.02.06.20019232

**Authors:** Jane Wray, Helen Gibson, David Barrett, Roger Watson

## Abstract

**Aim:** To undertake a rapid evidence assessment of approaches used to enhance nurse transition and retention for NQNs.

**Design:** A rapid evidence assessment.

**Data sources:** Electronic databases (CINAHL complete, Academic search premier, Open Grey, ERIC* (Education), Web of Science--Social Science Citation Index and PubMed)

**Review methods:** A rapid evidence assessment (REA) was undertaken to gain an overview of the density and quality of evidence on nurse transition and retention from student to Registered Nurse. Electronic databases were searched, and the full texts of relevant papers were retrieved and classified according to methodology. Studies were appraised using relevant CASP and MMAT tools and a single descriptor of quality: high; medium; or low was assigned to each output. Given the disparity in methods, the lack of randomized trials, results could not be combined; therefore, a descriptive approach was used to synthesise and present the data.

**Results:** Orientation and creating supportive environments were frequently reported as being effective in enhancing transition across a range of studies. A range of methods: quasi-experimental, survey and qualitative were used. Generally speaking the quality of most studies was poor.

**Conclusions:** Despite decades of research into the experiences of NQNs and development of schemes and frameworks to support them during this period, there is little substantive or robust evidence in terms of impact on retention. Further research into the longer-term retention of NQNs is recommended. Longitudinal studies would be beneficial in assessing the efficacy of approaches to enhancing retention.

**Impact:** Nurse managers need to work with education providers to facilitate experiences for final year nursing students to ease transition and also implement effective evidence based practices during the first year of registration and monitor the impact of this on retention.

## INTRODUCTION

Concerns regarding nursing workforce shortages and subsequent pressures on service delivery and patient experience remain prominent internationally (RCN 2018; The Health Foundation et al 2018; The Health Foundation 2017; Aiken et al 2014). In 2019 the UK Nursing and Midwifery Council (NMC) reported their register had grown by approximately 8,000 since 2018 (NMC 2019). Simultaneously however, NHS providers reported one of the most concerning issues facing them was ‘lack of nurses’ (NHS Providers 2019; 24). Furthermore, the number of applicants successfully entering undergraduate nursing courses in 2018 was 4% less than 2016 (NHS Providers 2019:24). Leaving the European Union and changes to how nursing students in the UK are supported financially (The Complete University Guide 2019) may have an impact. Nursing workforce demographics are changing. Recent analysis of NHS workforce trends by The Health Foundation (2019) suggest an increase in internationally recruited nurses and policy analysis suggests this needs to increase. International data on nursing shortages is not readily available because of differences between countries in how nursing shortages are measured (Drennan and Ross 2019).

### Background

The ageing of the nursing workforce is well documented (Buchan 1999; Graham and Duffield 2010; Kwok et al 2016; Buchan et al 2015), as is high staff turnover linked to stress, workload and burnout (Heinen et al 2013; Halter et al 2017). This has led to renewed interest on recruitment and retention of the existing nursing workforce. Strategic intervention is considered key to ensuring workforce stability and sustainability and this has led to many policies to focus employers on workforce retention (Health Education England 2015; The Health Foundation 2017; House of Commons Health Committee 2018; NHS Education for Scotland (NES) 2019).

Newly qualified nurses (NQNs) are one group considered at risk of leaving the workforce early. Reliable measures of this phenomenon internationally are not available as figures relating to this group of nurses tend to be conflated with turnover reports. Furthermore, the issue is clouded by semantics (Drennan and Ross 2019, Buchan et al 2018). The phrase ‘turnover’ is often used interchangeably with ‘attrition’ (Buchan et al 2018); a nurse leaving an organization might not necessarily leave the profession, they may move between jobs within the same organization. Estimated levels of attrition amongst NQNs vary. Up to 50% of nurses leave their first position within 1 year due to stressors in the work-place (Canada; Winfield et al 2009) and graduate nurse attrition rates of between 30 - 50% are reported in the USA (Phillips et al(2017). NQNs are thus a particularly vulnerable group and the challenge of transition from student to autonomous practitioner has been well documented in the literature and described as ‘reality shock’ (Kramer 1974) and ‘transition shock’ (Duchscher 2009).

Preceptorship is commonplace in many NHS trusts and private hospital settings in the UK and other schemes including mentorship and clinical supervision are considered fundamental to facilitating transition and developing safe and competent accountable practitioners (NMC 2006; Department of Health 2010). The Nursing and Midwifery Council (NMC) recommended:

> *all ‘new registrants’ have a period of preceptorship on commencing employment, this applies to those newly admitted to the NMC Register who have completed a pre-registration programme in the UK for the first time, or have subsequently entered a new part of the register. New registrants also include those newly admitted to the register from other European Economic Area States and other nation states* (NMC 2006:1).

NMC guidance recommends a formal period of preceptorship lasting “*about four months but this may vary according to individual need and local circumstances*” (NMC 2006: 2) and this is considered a ‘model of enhancement’ (NMC 2006; DH 2010) central to the continued professional development of the nurse rather than a framework to address educational deficits. The introduction of the UK Preceptorship Framework for Newly Registered Nurses, Midwives and Allied Health Professionals (Department of Health 2010) sought to reinvigorate preceptorship in the NHS and provide a commitment to staff and improved patient care. The DH guidance (2010) said that four months advocated in the NMC Guidance (2006) was insufficient.

Preceptorship may *‘include classroom teaching and attainment of role-specific competencies, however the most important element is the individualised support provided in practice by the preceptor ‘* (CAPITAL Nurse 2017:4). In the UK, whilst the preceptorship model is widely supported, the integrity of this approach is persistently compromised by lack of preceptors (Deasy et al. 2011; Whitehead et al. 2013, Adams and Gillman 2016). Workload pressures and staffing deficits further undermine opportunities for preceptorship support in NHS Settings (Lewis and McGowan 2015) and the existence of preceptorship frameworks is insufficient to ensure that this support is delivered in practice. Whilst preceptorship occurs post registration in the UK, this is provided for final year student nurses in the US and Canada (Robinson and Griffiths 2009; Currie and Watts 2012; Cubit and Ryan, 2011). Although there is agreement that such approaches have a positive impact on job satisfaction and intention to stay (Rush et al. 2013; Phillips et al. 2013; 2014), there is little evidence of impact on NQN retention figures in the UK, nor on patient outcomes (Robinson and Griffiths 2009).

Research focusing on NQNs and transition has focused on supportive frameworks, professional socialisation, learning development and competency (Adams and Gillman, 2016, Rejon and Watts 2014, Dinmohammadi et al 2013) which feature in policy guidance (Department of Health 2010) and previous reviews of transition experiences (Higgins et al. 2010; Rush et al. 2013; Whitehead et al. 2013; Murray-Parahi et al. 2016). Whitehead et al. (2013) were concerned with preceptorship and preceptors, whereas Rush et al. (2013) and Adams and Gillman (2016) explored different types of transition support. The link with retention is inferred or assumed rather than made explicit in the review criteria. Consequently, this review sought to identify robust primary research about transition and retention of NQNs.

### Methodology

A rapid evidence assessment (REA) is ‘a form of knowledge synthesis in which components of the systematic review process are simplified or omitted to produce information in a timely manner’ (Tricco 2015:2). The need to provide evidence-based recommendations in a time and resources efficiently has driven the development and widespread application of REAs (Varker et al 2015, Khangura et al 2012). Whilst systematic reviews are considered the ‘gold standard’ of knowledge synthesis and have become synonymous with ‘Cochrane Reviews ‘, the method is not without its drawbacks as these can be labour and time intensive and focused on a narrow set of research questions (Khangura et al 2012). REA methodologies comply with the rigour of the systematic review approach but in a shorter time with fewer questions (Thomas et al. 2013, Government Social Research Service (GSR) 2013).

Our research questions were:

- What approaches are used to enhance the transition of newly qualified nurses?
- What approaches are used to enhance retention of newly qualified nurses?
- What is the strength of the evidence for specific approaches to nurse transition and retention?

There is diversity within the literature about the length of time a NQN can still be considered a NQN or has achieved ‘competent status’ according to Benner (1984). For this review the term NQN is used throughout and defined as 12 months or less. This is consistent with earlier research exploring the stages of transition (Dearmun 2000, Evans 2001, Duchscher 2008, Andersson and Edberg 2010).

### Search Strategy

The search was undertaken using: specific database searching; and secondary searching of relevant websites. Electronic databases (CINAHL complete, Academic search premier, Open Grey, ERIC* (Education), Web of Science--Social Science Citation Index and PubMed) were searched during February 2018. The following search terms were used:

Newly qualified nurse **OR** Newly registered nurse **OR** New nurse **OR** Student Nurse **OR** Nursing Student

### AND

Transition **OR** Retention **OR** attrition **OR** Turnover **OR** stability Truncation, for example Nurs* for nurses, nurse and nursing and Boolean operators was also used to maximise search results. An inclusion and exclusion criteria was applied to focus the search approach and can be seen in Table 1.

**Table 1:**
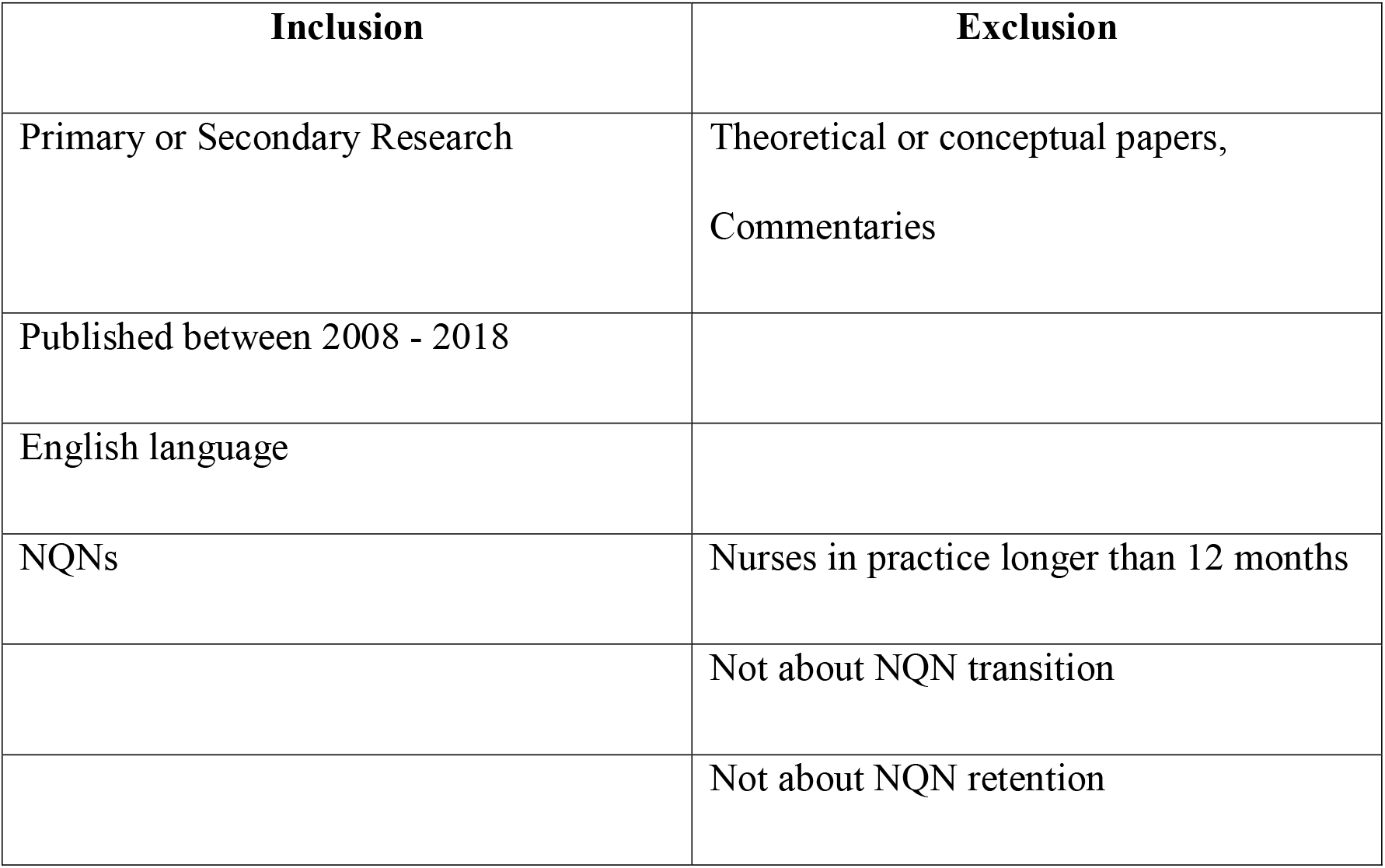
Inclusion and Exclusion Criteria

Papers were included if participants were NQNs, were primary research studies (defined as ‘*empirically observe a phenomenon at first hand, collecting, analysing or presenting ‘raw’ data ‘*) or secondary research studies, (defined as studies that: ‘*interrogate primary research studies, summarising and interrogating their data and findings* ‘) (https://www.gov.uk/government/collections/rapid-evidence-assessments; accessed 9 August 2019). Papers were included if published in English and between 2008–2018. The rationale for disregarding papers published before 2008 was that this date marked the publication of Duchscher ‘s (2008) seminal work on ‘transition shock ‘. Where papers examined NQN over a longer timeframe, they were only included if data were available to differentiate between those up to 12-month NQN and those beyond 12 months.

### Classification of Papers

Full texts were retrieved and classified using the RAE tool and template (https://www.gov.uk/government/collections/rapid-evidence-assessments; accessed 9 August 2019). Each member of the team (HG, DB, RW & JW) was randomly allocated papers for this preliminary classification. Papers were excluded at this stage if they did not meet the inclusion criteria.

### Quality Assessment

Each member of the team undertook quality assessment of the papers using an appropriate appraisal tool. CASP (Critical Appraisal Skills Programme) appraisal tools were used to assess qualitative papers, systematic reviews and RCTs. The MMAT (Mixed Methods Appraisal Tool) was used to assess mixed methods papers, and a customised tool based on Kelley et al. (2003) was use for evaluating surveys. Using a series of questions to prompt the reader to consider if the study was valid, what the results of the study were and if the results were useful these tools ensured a systematic assessment of the trustworthiness, relevance and results of published papers (https://casp-uk.net/). To ensure consistency, a subsample of the papers was quality assessed at this stage by another member of the team. Disagreement was resolved via discussion or involvement of a third team member.

### Final assessment and scoring

A single descriptor of quality: high; medium; or low was assigned to each output based on whether or not the output was low, medium or high in the range of scores afforded by the quality assessment instrument.

### Data Extraction

Key details of the included papers are summarised in Table 2.

**Table 2:** Summary of findings

| Study<br>Name (date) | Design<br>Experimental/survey<br>Qualitative/review | Results<br>Main findings | Study<br>quality &<br>Comments |
| --- | --- | --- | --- |
| Allan et al<br>(2018) | Qualitative | Findings suggest that the implementation of preceptorship can be highly variable. Inadequate formal preceptorship can be compensated for by informal on-ward support. Where there is neither sufficient formal preceptorship nor a lack of compensatory informal support, NQNs can struggle in putting their knowledge to work effectively and reworking it to meet the changing demands of ward-based practice | CASP:<br>Qualitative<br>LOW |
| Andersson and<br>Edberg 2010 | Qualitative | 2 key themes (1) up to 6 months - being a rookie (striving for acceptance, respect from colleagues), (2) from 6-12 months feeling like a registered nurse, (shoulder responsibility, prioritize tasks, and convey confidence to patients).<br>Being accepted and respected as a colleague was more important than being accepted and respected by patients for NQNs in first few months. | CASP:<br>Qualitative<br>LOW |
| Ashton 2015 | Correlational study was implemented | Being in a formal orientation period significantly supported the new | Survey<br>Assessment |
|  | using a cross-sectional design | nurses' overall adaptation. | Tool: High |
| Brandt et al 2017 | Qualitative | Themes<br>Pre-transition (1) A Combination of How the ASBSN Program Prepared Me and How I Prepared Myself. Orientation (3) - Intense Situations Evoked Strong Emotions; The Pace of Progression Fit My Needs. How I Spent My Time Was an Important Factor in Orientation<br>Post-orientation (3) Building My Own Support Network, Keeping the Patient Safe as I Built Confidence, Being on my Own Was Frightening<br>Being a NQN (2) I Had Supportive Colleagues and Plentiful Help and Experience Cultivated Confidence and I Grew Into a Contributing Team Member. | CASP:<br>Qualitative<br>LOW |
| Bratt & Feltzer 2012 | Longitudinal correlational design with data collected from 16 different cohorts of nurse residents over a three-year period from 2005-2008 | Initial job placement in the new graduate's desired position was found to be a significant predictor of organizational commitment; otherwise the results are very hard to ascertain | Survey Assessment Tool: Medium |
| Bridges et al 2013 | Qualitative | 3 themes<br>1 - becoming independent<br>2 - knowing the culture, 3 - relationship with the coach. | CASP:<br>Qualitative<br>LOW |
| Chandler 2012 | Qualitative | 3 themes<br>1 - They were there for me<br>2 - There are no stupid questions<br>3 - Nurturing the seeds | CASP:<br>Qualitative<br>LOW |
| Chappy et al 2010 | Qualitative | 1 major category (clinical instruction)<br>4 subthemes - more clinical time, more technical skills, broader range of real life experiences and communication with physicians. | CASP:<br>Qualitative<br>MEDIUM |
| Deasy et al 2011 | Not stated – but used survey method | Respondents had high levels of confidence in clinical abilities both at pre-registration and post-registration. They also perceived themselves to be competent across a range of other domains | Survey Assessment Tool: Low |
| Ebrahimi 2016 | Qualitative | 5 themes<br>1 - Lack of Support-Seeking Behaviors<br>2 - Management Weaknesses<br>3 - Ineffective Communication<br>4 - Personal Characteristics of Nursing Personnel<br>5 - Cultural Barriers | CASP:<br>Qualitative<br>HIGH |
| Ebrahimi et al (2016) | Qualitative | Emotional support emerged in four categories: 1)Assurance, 2)creating a sense of relaxation and security,3) lifting spirits, 4)emotional belonging and involvement | CASP:<br>Qualitative<br>HIGH |
| Edwards D et al (2015) | Narrative review | Findings suggest that transitional support arrangements such as internships, mentorship and preceptorship can have beneficial impact on competence, confidence, anxiety and job satisfaction amongst NQNs. There was some suggest that retention rates could be improved, though it was very difficult to link cause and effect. Interesting point made that doing something seems to be important, rather than worrying too much about what you do. | CASP:<br>Systematic Review<br>Medium |
| Gardiner and Sheen (2016) | Narrative review | Three themes:<br>Transition is a stressful experience; the need for a supportive | CASP:<br>Systematic Review |
|  |  | environment; importance constructive feedback during the transition to practice. | Medium |
| Gerrish 2000 | Qualitative – two cohort comparison (pre (1998) and post (1985) project 2000) | Three categories:<br>1 – “fumbling along”<br>2 – “taking charge of the ward”<br>3 – “metamorphosis”<br>Need for a bridging period over the latter part of the programme and first 6 months post – qualification to gradually acclimatize to accountable practitioner | CASP:<br>Qualitative<br>MEDIUM |
| Guay et al (2016) | Qualitative | In the early part of the transition, NGRNs experienced Surviving Without a Safety Net, which involved Experiencing Fear, Figuring It Out, and Learning on the Job. In the later part of transition, the NGRNs experienced Turning of the Tables, which involved Being Trusted, Gaining Confidence, and Feeling Comfortable in their professional role. | CASP:<br>Qualitative<br>MEDIUM |
| Harrison-White (2013) | Qualitative | Five themes emerged from the preceptees<br>1) Traumatic transition from student to staff nurse. 2) Added pressure of being a newly qualified children’s nurse 3) Need for proper accessibility and support from preceptors. 4) Need for a programme that is clinically focused. 5) View that preceptees should receive informal support outside of the formal guidelines of preceptorship. Five themes emerged from the Preceptors; 1) A requirement for formal establishment and structure of the preceptorship programme.<br>2) A goal of enabling preceptees to feel confident and comfortable in their new role and environment. 3) The importance of timely preceptorship at the beginning of the newly qualified nurses’ career journeys.<br>4) The importance of meaningful feedback.<br>5) Specific study on the programme should be ward-based, supported and relevant. | CASP:<br>Qualitative<br>LOW |
| Hollywood 2011 | Qualitative | Five primary themes (Paper only presents 2) (1) Support network (subtheme finding your own way, mentorship / preceptorship and previous experience) and (2) reality shock (subtheme professional accountability) | CASP:<br>Qualitative<br>MEDIUM |
| Hussein et al 2016 | Cross-sectional survey | Three independent and significant predictors of NGNs’ satisfaction were: (1) unit satisfaction; (2) satisfaction with the clinical supervision; and (3) assigned unit | Survey<br>Assessment<br>Tool: High |
| Johnstone et al (2008) | Qualitative | Three main findings: 1) support is critical to the process of graduate nurse transition, and that integration into “the system” is best provided during the first 4 weeks of a graduate nurse transition program and thereafter at the beginning of each ward rotation; 2) “informal teachers” and the graduate nurses themselves are often the best sources of support; 3) the most potent barriers to support being provided are the untoward attitudes of staff toward new graduates. | CASP:<br>Qualitative<br>MEDIUM |
| Ke Y T et al (2017) | Systematic review | Paper demonstrated that new nurses’ overall competence increased significantly due to preceptorship.<br>The most adopted preceptorship approach was a fixed preceptor/preceptee model and one-on-one for 1–3-month duration. | CASP:<br>Systematic<br>Review<br>High |
| Kuokkannen et al 2016 | A descriptive, cross-sectional and correlational design | Association between nurse empowerment and professional competence job satisfaction, and intent to change job, and other variables | Survey Assessment Tool: High |
| Labrague LJ and McEnroe-Petitte DM (2017) | Integrative review | Newly qualified nurses perceived low to moderate levels of stress, mainly as a result of heavy workloads and lack of professional nursing competence. | CASP: Systematic Review Medium |
| Lee et al 2009 | Quasi-experiment | Turnover 46.5% less than the previous year; cost saved US\$186,102; medication errors, falls and adverse effects all decreased | CASP RCT: Low |
| Lee et al (2013) | Qualitative | Transition process of new nurses becoming experienced members of the clinical nursing team was revealed as a journey of 'struggling to be an insider'. This phenomenon was characterised by four themes 1) 'being new as being weak', 2) 'masking myself', 3) 'internalising the unreasonable' and 4) 'transforming myself to get a position'. | CASP: Qualitative<br>MEDIUM |
| Lewis and McGowan (2015) | Qualitative | Two main categories emerged from the data.<br>1) Support Requirements which was further broken down into two themes: time and build confidence<br>2) Expectations of Preceptorship which was further broken down into two themes; understanding the process and understanding the preceptor's role. | CASP: Qualitative<br>MEDIUM |
| Linder (2009) | Qualitative | Eleven themes in the categories of professional role development, a unique practice, and personal reflection were identified. | CASP: Qualitative<br>MEDIUM |
| McDonald AW & Ward-Smith P (2012) | Narrative review | Papers suggested that the key elements of any orientation program are as follows: (a) evaluation of baseline knowledge, (b) inclusion of higher level skill practice, (c) support from an experienced individual (expert) in the unit where the new nurse will be working, (d) provision of opportunities to clarify existing knowledge and expand knowledge, and (e) evaluation of individual program outcomes. | CASP: Systematic Review<br>Low |
| Marks -Maran et al 2013 | Mixed Methods | Preceptorship was highly valued by the majority of preceptees (85%). Preceptors played a positive role in terms of alleviating stress. Preceptorship impacted positively on preceptees in terms of development of communication skills and clinical skills, and role, personal and professional development. In addition, preceptees felt that the programme was of value despite acknowledging difficulties in making time to meet with preceptors. Preceptees also indicated that they would wish to be preceptors in the future and that they would recommend preceptorship to all nurses who are either newly qualified or new in role. | MMAT:<br>75% (3 Stars) |
| Martin and Wilson (2011) | Qualitative | Two themes that are congruent with the literary and theoretical context within which the study is situated<br>1) Adapting to the Culture of Nursing<br>2) Development of My Professional Responsibilities | CASP: Qualitative<br>MEDIUM |
| Meyer et al | Two group comparative | 12% of control group and 6% of intervention changed jobs | CASP RCT: |
| 2017 | design over time | within 12 months; intervention group more likely to recommend nursing as a career at 3 months | Moderate |
| Mooney (2007) | Qualitative | Two categories, emerged 1) Learning the Ropes 2) The Metamorphosis | CASP:<br>Qualitative<br>MEDIUM |
| Moore and Cagle 2012 | Qualitative - interpretive study (Heideggerian phenomenology) | 3 key themes<br>1 - feeling the fit<br>2 - mentoring to push and pull growth<br>3 - proving competency: losing the apron strings. | CASP:<br>Qualitative<br>LOW |
| Muir et al (2013) | Mixed methods | Seven themes emerged. These were preceptors' perceptions of: the personal development of preceptees; the role development of preceptees; the communication skills development of preceptees; the clinical development of preceptees; the development of professional relationships by preceptees; value of the preceptorship programme to the organisation and value of being a preceptor in terms of their own professional development. | MMAT:<br>75% (3 Stars) |
| Olejniczak EA et al (2010) | Integrative review | Three themes were identified: socialization to the professional role, competence and confidence in self-performance, and learning in a safe and supportive environment | CASP:<br>Systematic Review<br>Low |
| Oosterbroek TA (?date – certainly post 2015) | Integrative review | Four main themes emerged:<br>The nature of the rural experience, the importance of interprofessional collaboration, issues related to recruitment and retention of nurses to rural communities, and factors associated with student performance evaluation and feedback | CASP:<br>Systematic Review<br>Low |
| O'Shea and Kelly (2007) | Qualitative | Two themes emerged from the data;<br>1) The experience of being qualified: highs and lows<br>2) Stressful aspects of the staff nurse role. | CASP:<br>Qualitative<br>MEDIUM |
| Pasila K et al (2017) | Integrative review | Newly graduated nurses' had four categories of orientation experience:<br>Experiences related to orientation arrangements;<br>Experiences related to the preceptor;<br>Experiencing role transition during the orientation; | CASP:<br>Systematic Review<br>Medium |
|  |  | Suggestions for changes based on orientation experiences.<br>It was found that the orientation and the preceptor have a great impact on how newly graduated nurses experience the start of their career. |  |
| Penprase 2012 | Qualitative | 2 key areas<br>1 - aspects of the students' educational experience that prepared them for the professional nursing role<br>2 - perceptions and factors contributing to the participants' satisfaction with their current nursing | CASP:<br>Qualitative<br>LOW |
| Phillips et al 2012 | A descriptive questionnaire survey | Transition scores were significantly higher for undergraduates who were employed; institutional work factors appeared to be stronger predictors of successful transition than pre-registration employment factors. Assistance in dealing with complex patients, orientation to a new environment, and respect from colleagues were the best predictors for successful transition | Survey<br>Assessment<br>Tool: High |
| Roziers and Ramugondo (2014) | Qualitative | Three general themes emerged 1) a sense of achievement; 2) uncertainty and fear in anticipation of reality 3) reality shock | CASP:<br>Qualitative<br>MEDIUM |
| Spiva, L et al 2013 | Qualitative | Four major themes or patterns (10 sub themes)<br>1 - Preceptor variability 2 - Professional growth and confidence changed with time<br>3 - A sense of being nurtured<br>4 - Enhancements needed to improve orientation experience | CASP:<br>Qualitative<br>LOW |
| Steen et al 2011 | Not stated – but survey design | The programme eased transition | Survey<br>Assessment<br>Tool: Low |
| Tseng et al 2013 | Pretest-posttest quasi experiment | Multiple outcomes including the fact that nurses in the intervention group were more likely to be in their first hospital after 1 year | CASP RCT:<br>Moderate |
| Walker et al (2013) | Qualitative | GNs and NUMs differed with respect to perceptions of unprofessional workplace behaviour and coping with death and dying. The findings suggest that NUMs and GNs do not always have a shared understanding of the stressors that GNs face in the first year of clinical practice | CASP:<br>Qualitative<br>MEDIUM |
| Walton et al (2018) | Qualitative | Five key themes were identified. The students' reflections noted individual attributes - personal and professional strengths and weaknesses; professional behaviour - actions such as engaging help and support, advocating for patients' needs and safety and putting their own feelings aside; situational challenges such as communication difficulties, both systemic and interpersonal, and the pressure of competing demands. Students also identified rewards - results they experienced such as achieving the nursing outcomes they desired, and commented on reflection as a useful tool. | CASP:<br>Qualitative<br>MEDIUM |
| Ward A & McComb S (2017) | Narrative review | Findings were from both the preceptor and preceptee perspective.<br>Themes related to preceptor perspective were satisfaction and confidence, support, role preparation, and workload issues.<br>Preceptee perceptions were categorised under satisfaction and confidence, support, and preceptor effectiveness. | CASP:<br>Systematic<br>Review<br>Low |
| Watt and Pascoe (2013) | Qualitative | Three major themes emerged;<br>1) Being situated in a clinical school within the hospital<br>2) The university away from the university<br>3) Engagement with practice | CASP:<br>Qualitative<br>LOW |
| Ya-Ting Ke and Min-Tao Hsu 2015 | Qualitative | 3 major themes:<br>1 - the stages of a preceptorship<br>2 - the differences of preceptorship<br>3 - quasi-family functions. | CASP:<br>Qualitative<br>MEDIUM |
| Yeh & Yu 2009 | A cross-sectional research design | Intention-to-quit group had significantly higher job; not having practiced in the working hospital was associated with the intention to quit | Survey<br>Assessment<br>Tool: High |

## RESULTS

References were imported into Endnote and duplicates removed leaving 2,647 across the databases (Table 3). Two further titles were identified via web searches. Titles of the 2,649 references were examined for relevance; 106 were excluded as they were not primary or secondary research. A further 2,239 not meeting the inclusion criteria were excluded (Table1). A further 123 papers were removed after examining the abstracts

**Table 3:** Number of references per database

| Database Name | Number of References |
| --- | --- |
| Academic Search Premier | 838 |
| CINAHL | 1099 |
| ERIC | 30 |
| PubMed | 237 |
| Web of Science | 2065 |
| Subtotal | 4269 |
| Duplicates | 1622 |
| **Total** | **2,647** |

Full texts of the remaining 181 papers were examined and classified using the REA tool and template (https://www.gov.uk/government/collections/rapid-evidence-assessments). Each member of the team was randomly allocated papers for this preliminary classification. A further 133 were excluded because they did not meet the inclusion criteria (n=44 were not reporting research, n = 49 were not about NQNs, n = 11 were not related to transition and/or retention (n=11) and n = 10 for ‘other’ reasons, for example, the authors did not define a NQN). A further 19 papers were excluded because they were considered too low quality leaving 48 papers for data extraction; RCTS (3), Survey / other quantitative methodologies (8), systematic Reviews (8), qualitative studies (27) and mixed methods studies (2). The included papers were from the USA (n=17), the UK (n=11) Australia (n=8), Taiwan (n=3), Canada (n=3), Singapore, Finland, New Zealand, Sweden, Iran and South Africa (all n=1).

### Experimental or quasi experimental studies / RCT ‘s

All experimental studies (Lee et al 2009; Meyer et al 2017; Tseng et al 2013) were pre-post intervention so most CASP criteria did not apply and none had blinding of participants. Lee et al (2009) showed a reduction in turnover as a result of a preceptorship programme and provided a cost analysis and the effect on medication errors; both were reduced. Meyer et al (2017) and Tseng at al (2013) both showed reduced job changing in the intervention group which had experienced a revised curriculum before qualifying (Meyer et al 2017) and a practicum at the end of their courses was based on collaboration between their schools and their hospitals and a jointly developed programme (Tseng et al 2013).

### Quantitative studies

Quantitative studies predominantly used a survey design. They described a range of outcomes related to the study of the successful employment of NQNs and all were proxies for retention, including: Adaptation (Ashton 2015); Organisational commitment (Bratt 2012); Confidence and competence (Deasy 2011); Satisfaction with practice; Empowerment and competence (Kuokannen 2016); Predictors of successful transition (Phillips 2012); Effect of a programme of transition (Steen 2011); and Stressors and intention to quit (Yeh 2009).

All studies reported at least one positive outcome related to the aim(s) of the study, and from the highest quality studies it appears that effective strategies with a positive influence on proxy measures of successful NQN employment include: having a formal orientation period (Ashton 2015); the initial placement (Bratt 2012; Hussien 2016); satisfaction with the unit and clinical supervision (Hussein 2016); empowerment (Kuokannen 2016); pre-registration employment (Phillips 2012); and higher stress and not having had previous experience in the unit (Yeh 2009). The study by Deasy (2011) did not investigate the effect of independent variables and made no comparison with reference values of the levels of proxy measurement they used: confidence and competence (Deasy 2011).

### Systematic Reviews

Labrague & McEnroe-Petitte (2017) explored the experiences of new nurses during transition, and Ke et al, (2017), Oosterbroek (2017) and Ward & McComb (2017) focused on preceptorship. Olejniczak et al (2010) provided an overview of the evidence base linked to simulation as an orientation strategy for NQNs, whilst three reviews (Edwards et al (2015), Gardiner & Sheen (2016) and Pasila et al (2017) examined broader supportive approaches.

Where experiences of NQNs were discussed as part of a review, it was clear that transition from student to Registered Nurse is stressful (Labrague & McEnroe-Petitte, 2017; Gardiner & Sheen, 2016). Labrague & McEnroe-Petitte (2017) identified the main causes of stress were workload and a perceived lack of competence.

The need for a supportive environment providing NQNs with learning opportunities and constructive feedback was identified by Gardiner & Sheen (2016), Oosterbroek (2017) and Olejniczak (2010) who also found simulation could play an role in orientating new nurses, allowing for learning in a safe environment.

Benefits of preceptorship were discussed by Ward & McComb (2017), Ke et al (2017), Oosterbroek (2017), and Pasila et al (2017). Edwards et al (2015) explored the impact of a broader range of interventions: preceptorship; mentorship; and internship. These included the support offered by an experienced member of staff, and the provision of teaching and feedback. Only Ke et al (2017) evaluated specific approaches to preceptorship and reached firm conclusions that a fixed preceptor/preceptee model with regular one-to-one working was the most prevalent approach; preceptorship significantly increased NQNs competence, though no firm conclusions could be reached regarding impact on retention.

### Qualitative studies

methods included participant observation; interviews (both unstructured and semi-structured) and focus groups. The studies by Chappy et al (2010) and Penphase (2012) used text-based (qualitative) responses from a larger survey and Walton et al (2018) analysed ‘reflective essays’ written. Samples varied from four (Bridges et al 2013) to 94 (Walker et al 2013). Penphase (2012) reported 460 however these were text-based comments in a larger survey.

Analysis was generally well described with most frequently used Thematic analysis (n=7) and Content analysis (n=7). The constant comparative method, associated with grounded theory was also used (Guay et al 2016; Gerrish 2000; LeeAnna Spiva et al 2013). Linder (2009) and Watt and Pascoe (2013) did not give a specific name to their approach, however within each paper the process of data analysis was sufficiently described. Penprase (2012) provided detailed findings without indication of the method used or how data were selected.

Some studies looked at transition experiences then characterised these experiences into proposed ‘stages’ (Anderson and Edberg 2010 – 2 stages, Gerrish 2000 – 3 stages) i.e. driving theory, or the characteristics of the phenomena of the NQN (Moore and Cagle 2012). Whereas Brandt et al (2017) used existing frameworks for transition and focused on experiences that characterised particular stages (e.g. preparation or pre-transition, orientation/induction, post-orientation and being a NQN). Allan et al (2018), Harrison-White (2013), Lewis and McGowan (2015), LeeSpiva et al (2013) and Ya-Ting Ke and Min-Tao Hsu (2015) focused mainly on preceptorship

Support featured to some extent in all the papers; the importance of positive and supporting experiences/environments (Chandler 2012) and the impact on NQNs of being unsupported during this period (Hollywood 2011 ‘finding your own way ‘, Gerrish 2000 ‘fumbling along ‘). Key to ‘positive’ support experiences was acceptance by colleagues/the team and becoming a team member (Anderson and Edberg 2010, Brandt et al 2017, Moore and Cagle 2012).

Educational preparation for transition to practice featured in Brandt et al (2017), Chappy et al (2010), Penphrase (2012) and transition shock (a key feature of a much earlier study in the field) featured only in Hollywood (2011). Papers that explored becoming an accountable practitioner focused on growth and maturation during the later stages of transition (Anderson and Edberg 2010, Brandt et al 2017, Bridges et al 2013, Gerrish 2000). Satisfaction with current role in nursing was only explored by Penphase (2012).

### Mixed Methods studies

Two papers which reported on different aspects of the same study used mixed methods. The study was an evaluation of a preceptorship programme for NQNs in one NHS Trust. Marks-Maran (2013) presented data from the perspective of the preceptees and Muir et al (2013) presented data from the perspective of the preceptors. Quantitative data were collected using questionnaires and qualitative data were collected using reflective journals, reflective recordings and interviews. Marks-Maran et al (2013) found that preceptorship was highly valued by most preceptees (85%) with engagement in the programme perceived as having a positive impact on stress levels, role and personal and developing professional skills. Muir et al (2013) found the preceptorship progamme was viewed positively by preceptors and impacted positively on development of preceptees and development of preceptors themselves. The preceptorship programme was also viewed as of long-term benefit to the organisation.

## DISCUSSION

### What approaches are used to enhance the transition of newly qualified nurses?

The benefits of preceptorship were discussed in a number of papers including systematic reviews by Ward & McComb (2017), Ke et al (2017), Oosterbroek (2017), and Pasila et al (2017). However, only one paper (Ke et al 2017) evaluated specific approaches to preceptorship. The authors concluded that a fixed preceptor/preceptee model with regular one-to-one working was the most prevalent approach and this model of preceptorship significantly increased NQNs competence.

A recurrent theme within the papers, which may be considered a less formal approach to supporting transition, was the existence of a supportive organisational culture which may include being accepted by team and, or peers, effective communication within and across an organisation and access to and availability of informal support (peers, friends, the wider MDT).

Although firm conclusions on the impact of interventions were hindered by methodological weaknesses of the primary research in the review by Edwards et al (2015), most interventions resulted in some level of benefit related to competence, confidence and job satisfaction. However, this seemed less dependent on the type of intervention and was more broadly related to organisations demonstrating their commitment to supporting NQNs. Simply, the fact that something was being done was more important than the specific nature of the intervention.The only RCT paper that explored transition was Meyer and this related to curriculum changes.

### What approaches are used to enhance retention of newly qualified nurses?

Five of the studies directly addressed retention. Lee et al (2009) showed a reduction in turnover as a result of a preceptorship programme and provided a cost analysis of the effect on medication errors, both of which were reduced. Yeh et al (2009) found that higher levels of stress influenced intention to quit and Ya Ting et al (2015) found that having a good relationship with a preceptor positively influenced NQNs intention to stay. Two further papers McDonald and Ward Smith (2012) and Ke (2017), both literature reviews, looked at retention. McDonald and Ward-Smith (2012) concluded that longer interventions increased retention however the paper was evaluated as being of low quality so findings should be treated with some caution. Ke (2017) failed to reach any firm conclusions regarding the impact on retention rates of various models of preceptorship.

However, in most studies, retention was more likely to be inferred where transition experiences were positive. Thus, if we are to assume that positive transition experiences impact positively on retention rates the solution to retaining NQNs might be found in the evidence for what constitutes a good NQN transition programme. The qualitative studies found that indicators of successful transition (and implied enhanced retention) included formal organisational enablers; induction, orientation and supporting framework (mentoring, preceptorship, clinical coach), supportive organisational culture (acceptance by team/peers, effective communication within and across organisation) and access to and availability of informal support (peers, friends, the wider MDT/units). These enablers were seen as increasing job satisfaction (and impact on retention) and vice versa. Quantitative studies suggest that effective strategies that have a positive influence on proxy measures of successful NQN employment include: having a formal orientation period (Ashton 2015); the initial placement (Bratt 2012; Hussien 2016); satisfaction with the unit and clinical supervision (Hussein 2016); empowerment (Kuokannen 2016); pre-registration employment (Phillips 2012); and higher stress and not having had previous experience in the unit (Yeh 2009).

### What is the strength of the evidence for specific approaches to nurse transition and retention?

The quality of papers across the five methodological categories was variable. The quality of the three experimental studies (Lee et al 2009; Meyer et al 2017; Tseng et al 2013) retrieved was low or moderate with a particular weakness being a lack of blinding of participants. The quality of the quantitative studies included in the review varied from low (Deasy et al 2011) to high with only one study (Bratt & Feltzer 2012) rated medium. Deasy et al (2011) did not define the design of their study and Bratt and Feltzer (2012) provided no information about response rate to their survey. The remaining five studies using survey methods provided strong evidence, on the basis of the evaluation instrument designed for this study.

Overall, the standard of systematic reviews could have been higher. Only three of nine studies provided the necessary level of insight into search methodologies, critical appraisal and analysis of evidence. Though all systematic reviews used established databases as part of their literature search, the detail provided of methods, inclusion criteria and search terms varied substantially. Papers such as Edwards et al (2015), Ke et al (2017) and Pasila et al (2017) – which were judged to be the highest quality - provided a flowchart summary of the literature selection process in line with PRISMA (or similar) guidelines. In other papers, the account of the literature search was lacking in detail, thereby reducing confidence in whether all relevant evidence was identified. Edwards et al (2015), Ke et al (2017) and Pasila et al (2017) provided insight into the critical appraisal of selected papers, such as the use of Joanna Briggs Institute checklists. Some other papers highlighted methodological issues in the selected papers (though they provided no details on how these conclusions were reached). The lowest quality papers – Oosterbroek (2017); Ward & McComb (2017); Olejniczak (2010) – offered no discussion of how literature was appraised, analysed or quality assured. Some of the reviews included quantitative research, but none of them performed meta-analysis of data. All eight reviews offered a narrative of papers, with some identifying core themes and/or articulating the commonalities and areas of disagreement within the selected papers.

Similarly, there was considerable variation in the qualitative studies regarding reporting styles with quality ranging from very poor (met few of the CASP criteria) to moderate to high (met at least half or most of the CASP criteria). No studies met all the CASP criteria. The higher-quality papers were Ebrahimi (2016), Chappy et al (2010) Gerrish (2000), Hollywood (2011), Ya-Ting Ke and Min-Tao Hsu (2015), Ebrahimi et al (2016) and Linder (2009) and the lower quality studies were by Penprase (2012), Bridges et al (2013), Allen et al (2018) and Harrison-White (2013). The relationship between the researcher and participant and impact of potential bias and influence – key to qualitative methodologies was the most under-reported item across all the studies, followed by rationale for study design.

Both mixed methods studies were evaluated as being of good quality, scoring 75% (three out of four stars) using the MMAT. However, both papers failed to report on how the findings relate to the researchers’ influence and on the limitations of integrating qualitative and quantitative data both of which are methodological quality criteria within the MMAT.

## Limitations of the review

Limitations to our review should be noted. This was a rapid evidence assessment therefore some compromises were made (Thomas et al. 2013). In the types of studies retrieved, we only considered articles published in the past 10 years. Whilst we included studies from any country, the range of countries from which the studies in this review are included limits the ability to draw any conclusions that may be applied internationally as differences between the healthcare systems of different countries and the cultural and educational systems, mediates the experiences of NQNs.

Our definition of NQNs may have limited the articles retrieved and the timespan of 12-months after qualification may also have limited our search. There is a distinct lack of standardisation in relation to the definition of NQN in the literature which warrants further investigation. Further research into the longer-term retention of NQNs is recommended. Longitudinal studies would be beneficial in assessing the efficacy of approaches to enhancing retention and as such our findings relate to short-term retention (that is, in line with our definition of NQNs as being within the first 12 months of practice). Notwithstanding these limitations, this review has addressed a key gap in the available evidence relating to the density and quality of evidence on nurse transition and retention from student to registered nurse.

## CONCLUSION

A range of methods, varying in quality, has been applied in the past decade to the study of the retention of NQNs and those methods have been applied to a wide range of initiatives and programmes designed to increase such retention. However, the lack of a standard definition of ‘retention’ should be borne in mind when interpreting our findings. Therefore, it is hard to draw firm conclusions about the best ways to achieve retention. Nevertheless, orientation, and creating supportive environments were frequently reported as being effective across a range of studies. In terms of the hierarchy of evidence (Murad et al 2016), with systematic reviews the top and followed by randomised controlled trials, the overall assessment of the strength of evidence in this review is low. Systematic reviews were of poor quality, and experimental studies were all quasi-experiments. With no studies being large scale or long term, there is a definite need for longer term assessment of the impact of interventions. Surveys studies were, generally, of the highest quality, but they tended to use proxy measures of retention and in such correlative designs the relationship between cause and effect is weak.

## Data Availability

This is a review - no data as such available.

